# Gut Microbiota and Parasite Dynamics in an Amazonian Community Undergoing Urbanization in Colombia

**DOI:** 10.1101/2025.04.16.25325921

**Authors:** Sebastián Díaz, Amie J. Eisfeld, Mónica Palma-Cuero, Nathalie Dinguirard, Leah A. Owens, Karl A. Ciuoderis, Laura S. Pérez-Restrepo, John D. Chan, Tony L. Goldberg, Jessica L. Hite, Juan Pablo Hernandez-Ortiz, Yoshihiro Kawaoka, Mostafa Zamanian, Jorge E. Osorio

## Abstract

Studies on human gut microbiota have recently highlighted a significant decline in bacterial diversity associated with urbanization, driven by shifts toward processed diets, increased antibiotic usage, and improved sanitation practices. This phenomenon has been largely overlooked in the Colombian Amazon, despite rapid urbanization in the region. In this study, we investigate the composition of gut bacterial microbiota and intestinal protozoa and soil-transmitted helminths (STHs) in both urban and rural areas of Leticia, which is located in the southern Colombian Amazon. Despite their geographic proximity, the urban population is predominantly non-indigenous, while indigenous communities mostly inhabit the rural area, resulting in notable lifestyle differences between the two settings. Our analyses reveal a reduction in bacterial families linked to non-processed diets, such as *Lachnospiraceae, Spirochaetaceae*, and *Succinivibrionaceae*, in the urban environment compared to their rural counterparts. Interestingly, *Prevotellaceae*, typically associated with non-processed food consumption, shows a significantly higher abundance in urban Leticia. STH infections were primarily detected in rural Leticia, while intestinal protozoa were ubiquitous in both rural and urban areas. Both types of parasites were associated with higher gut bacterial richness and diversity. Additionally, microbial metabolic prediction analysis indicated differences in pathways related to unsaturated fatty acid production and aerobic respiration between rural and urban bacterial microbiomes. This finding suggests a tendency towards dysbiosis in the urban microbiota, possibly increasing susceptibility to non-communicable chronic diseases. These findings provide new insights into the impact of urbanization on gut microbiota dynamics in the Amazonian context and underscore the need for further research to elucidate any associated health outcomes.

**Relevance:** Changes in the diversity and composition of gut microbiota in urban populations have been linked to the rise of non-communicable chronic diseases, such as autoimmune conditions, diabetes, and cancer. As developing countries undergo a demographic shift towards increased urbanization, accompanied by changes in diet, housing, and medication use, there is a concerning loss of microbial diversity. Therefore, it is essential to investigate microbiota changes in overlooked populations, such as indigenous communities in the Colombian Amazon basin. A better understanding of local and generalizable changes in gut microbial composition through urbanization may facilitate the development of targeted programs aimed at promoting lifestyle and diet changes, to prevent diseases that healthcare systems may be ill-equipped to effectively address.

## Introduction

The taxonomic composition of the human gut microbiota is dynamic and shaped by factors such as diet, medication, sanitation, and occupational exposures [1]. Surveys on microbiota composition during population rural-to-urban transition have consistently observed a reduction in microbial diversity, particularly of bacterial groups associated with traditional lifestyles. These are known as VANISH (volatile and/or negatively associated with industrialized human societies) taxa. Examples include *Prevotellaceae, Spirochaetaceae*, and *Succinivibrionaceae* [2–5], whose decline is attributed to dietary shifts towards processed foods, widespread antibiotic usage, and decreased physical and outdoor activity. This is accompanied by a corresponding expansion of BloSSUM (bloom or selected in societies of urbanization/modernization) taxa, such as *Bacteroidaceae* and *Verrucomicrobia*, which are linked to an increased incidence of chronic disease [6,7]. These trends are expected to increase with the global rise of urbanization, with over half of the world’s population currently residing in urban areas [8].

Gut bacteria also interact with commensal and parasitic eukaryotes in the human host, including protists and soil-transmitted helminths (STHs) [9,10]. These single-celled and multicellular organisms interact with the bacterial community and are associated with increased richness and diversity, or changes in the abundance of specific bacterial taxa [11–17]. For example, abundance of the *Prevotella* genus of gut bacteria is positively associated with *Blastocystis spp*. and *Endolimax nana* protozoa [12], but negatively associated with the presence of *Entamoeba* [18]. These interactions can also be clinically relevant, such as the negative correlation between the *Megasphaera* genus of bacteria and diarrheal symptoms during *Cryptosporidium* infection [19]. Helminth parasites also alter host-microbiota interactions [20], with several well-known immunomodulatory effects of helminth infection requiring host microbiota [21,22]. Infection with *Trichuris muris* and *Heligmosomoides polygyrus* roundworms inhibits proinflammatory bacterial taxa while promoting colonization with protective *Clostridiales* species, and this relationship can be reversed with deworming [23]. The relative abundance of *Clostridiales* also changes in individuals infected with hookworm (*Necator americanus*) following anthelmintic treatment [24]. Both protozoa and helminth parasites also stimulate intestinal tuft cells [25,26]. These cells promote type II immunity and alter intestinal microbiota composition [27,28] while also secreting signals that directly modulate parasite biology [29].

Demographic and cultural change in the Amazonian region is characterized by an urban expansion of existing riverside towns and the establishment of new peri-urban settlements driven by agriculture, extractive industries, and infrastructure development [30]. However, gut microbiota studies have predominantly focused on rural horticulturists and hunter-gatherers, with limited attention given to urban populations [31–36]. Leticia, situated in the southern Colombian Amazon, serves as the capital city of the Departamento del Amazonas and forms an urban complex straddling the borders of Brazil, Colombia, and Peru. The city’s population is estimated at 100,000 inhabitants, with distinct demographic characteristics observed between urban and peri-urban rural areas [37,38]. Mixed-race populations predominantly inhabit the urban area, while indigenous groups primarily populate rural communities settled in response to extractive booms (e.g. rubber and coca) and assimilation efforts such as missionary campaigns [39].

This study provides novel insights into the urban and rural gut microbiota and parasite communities of a Colombian Amazonian population. We aim to understand how social factors, medical history, and current infection with parasitic protozoa and STHs interact with the bacterial microbiota composition. Additionally, using metabolic pathways predictions, we explore the potential health implications of the bacterial taxonomic differences between urban and rural areas.

## Materials and methods

### Ethics and Human Subjects

All work conducted in this study received approval from the Research Ethics Committee of Universidad Nacional de Colombia (protocol number CEMED-060-19), the University of Wisconsin-Madison Health Sciences Institutional Review Board (protocol number 2020-0214), and local indigenous community leaders. Healthy volunteers were recruited in March 2021 from two field sites: (a) the peri-urban rural multi-ethnic indigenous community of Nimaira Naimeki Ibiri Kilómetro 11 (referred to as Km11) (n = 80); and (b) within the urban city limits of Leticia (referred to as Leticia) (n = 20). Colombian Army and Air Force support facilitated operations at the Km11 field site. Before enrollment, consent was obtained from all participants. For individuals under 18 years old, formal written consent was provided by their parents or legal guardians. Before sampling, each participant completed a survey addressing socioeconomic and health status.

### Donor sampling

Enrolled participants were provided with a specialized kit for fecal specimen collection, along with verbal instructions for proper sample acquisition. Upon collection, fecal specimens were promptly stored in pack ice for transportation to the State Public Health Laboratory. Aliquots of 1 g of the specimens were preserved in 2 ml of DNA/RNA Shield (Zymo Research) and stored at −80°C. Preserved specimens were transferred to the One Health Genomic Laboratory (OHGL) at the Universidad Nacional Sede Medellín and shipped to the University of Wisconsin-Madison for DNA extraction and sequencing. In addition to fecal specimens, serum samples and nasal swabs were collected for SARS-CoV-2 testing. Recent infections were assessed using the Abbott Architect SARS-CoV-2 IgG antibody assay in serum (Abbott Park, IL). Active infections were determined through genomic DNA extraction from nasal swabs using Gene E RT-PCR, as previously described [40].

### DNA Extraction

DNA extraction from fecal specimens was conducted using the QIAamp PowerFecal Pro DNA kit (Qiagen). The extraction process followed the manufacturer’s protocol, with a modification at Step 1, where the input material consisted of 50 μl of fecal slurry in DNA/RNA Shield, 500 μl of CD1 buffer, and 300 μl of ATL buffer (Qiagen; not included in the kit). Bead beating was performed using a TissueLyser II (Qiagen) for two cycles, each lasting 5 minutes at 25 Hz. Between cycles, adaptors containing the specimens were repositioned so that samples that were closer to the machine body were further away in the second cycle. Finally, samples were eluted in a final volume of 50 μl RNase-free water and stored at −80°C.

### 16s rRNA metabarcoding sequencing and analysis

Qiagen Genomic Services conducted 16S rRNA microbiome profiling using the QIAseq 16S/ITS Screening Panel for library preparation. First, starting with 1 ng of DNA, target regions were selected and amplified through 12 cycles of PCR. Samples underwent cleanup using QIAseq Beads (Qiagen), followed by the addition of sequencing adapters and enrichment in a second PCR of 12 cycles. After a second bead cleanup, the libraries underwent quality control assessment using capillary electrophoresis (Tape D1000). High-quality libraries were then pooled in equimolar concentrations, determined by the Bioanalyzer automated electrophoresis system (Agilent Technologies). The library pool(s) were quantified using qPCR, and the optimal concentration was used to generate clusters on the surface of a flow cell before sequencing on a MiSeq (Illumina Inc.) instrument (2×276). Sequencing data were deposited in the NCBI SRA database (bioproject accession number PRJNA1246579).

Raw V4-V5 16S rRNA fragment reads were processed using a QIIME2 pipeline [41]. The DADA2 plugin [42] was utilized for trimming reads, removing sequences with ambiguous nucleotides and chimeras, and discarding singletons. The remaining sequences, with a length of approximately 370 bp, were clustered into Operational Taxonomic Units (OTUs) at a 99% identity level. Taxonomic classification was performed using the q2-feature-classifier plugin with the Bayes machine-learning classifier method [43] trained with the Greengenes 515F/806R database v.13.8 [44] with OTUs identified as Mitochondria, Chloroplast, or Archaea discarded. After removing low abundance OTUs (≤ 0.01% total sampling) and samples (≤ 1000 sequences), two individuals from Km11 were discarded, resulting in a final sample size of 98 individuals.

Alpha and beta diversity analyses were conducted in R in the phyloseq package [45]. To assess the overall influence of donor location, a permutational multivariate analysis of variance (PERMANOVA) with 999 permutations was performed on weighted UniFrac distances using the vegan package [46]. To identify microbial composition differences between the locations, two approaches were employed: (a) identify OTUs with differential abundance using the Wald significance test implemented in DESeq2 [47], with Km11 as the treatment group and Leticia as the control (threshold cutoff, α = 0.01); (b) evaluate the differential abundance of selected BloSSUM and VANISH taxa by using Wilcoxon paired tests. For BloSSUM taxa, *Bacteroidaceae*, *Verrucomicrobiaceae*, and *Rikenellaceae*, were chosen. For for the VANISH taxa, *Prevotellaceae* plus *Paraprevotellaceae* (referred as *Prevotellaceae*), *Succinivibrionaceae*, *Spirochaetaceae*, and the clostridiales *Lachnospiraceae* with *Ruminococcaceae* as one group (referred as *Lachnospiraceae*) were chosen. For microbiota functional analysis, PICRUSt 2.0 [48] was utilized to predict biological pathways. Results were classified with the MetaCyc database [49] and differential abundances between Km11 and Leticia microbial pathways were evaluated using a Wald significance test within the ggpicrust2 package [50].

To describe the Leticia bacterial microbiota structure in a regional context, we performed a comparative analysis of our dataset against previously reported Amazonian and non-Amazonian Colombian microbiotas. We included datasets that meet two criteria: used the V4 region of the 16S rRNA sequence and have reads with >100 bp in length. The final analysis included nine datasets, divided into six Amazonian populations: (a) the urban and rural Leticia sampling as one group; (b) the urban Belém and rural indigenous (c) Suruí, (d) Tupaiú, and (e) Xikrin [4] communities in Brazil; and the (f) rural indigenous Tsimané community in Bolivia [34]; and three non-Amazonian datasets, two urban Colombian (g) Bogotá and (h) Medellín) [45]; and (i) an urban American cohort from Ohio, subsampled from the American Gut Project [46] **(Table S1)**. Given that some datasets only sampled adult donors, infant samples (<15 years old) were removed before the analysis. Raw 16S rRNA reads were trimmed to 130 bp and preprocessed using the QIIME2 pipeline. Comparative analysis of alpha and beta diversity and selected BloSSUM and VANISH taxon abundance were performed in R.

### 18s rRNA metabarcoding sequencing and analysis

For the same DNA samples used for 16s rRNA analysis, we carried out eukaryotic analysis using the VESPA (Vertebrate Eukaryotic Endo-Symbiont and Parasite Analysis) metabarcoding protocol [51] targeting the 18S rRNA gene V4 region. Library pools were sequenced using MiSeq (Illumina Inc.) instrument with a 300×300 cycle chemistry. Sequencing data were deposited in the NCBI SRA database (bioproject accession number PRJNA1246579). Raw reads were processed using a QIIME2 pipeline with OTUs at a 99% identity level classified using the PR2 reference sequence database v.5.0 [52]. OTUs with unassigned or incomplete taxonomy using the PR2 database were manually classified using the full NCBI nucleotide database. Three Km11 samples with ≤ 1000 sequences were discarded from the final dataset. PERMANOVA tests were used to evaluate the influence of parasitic protists and nematodes in bacterial community structure.

## Results

We evaluate the impact of urbanization on local microbiota structure in the gut microbiota of Leticia by comparing the peri-urban rural indigenous community of Kilometro 11 (Km11) with the non-indigenous urban population (Leticia). Participants completed surveys that provided insights into the conditions of the community (**Table 1**). The cohort showed a representative sex distribution (Leticia, 60% female, 40% male; Km11, 58% female, 42% male) with donors ranging from 5 to 82 years (Leticia, median age 36.5 years old; Km11 38.0 years old). As expected, ethnic identification varied significantly by location, with 95% of the Km11 community identifying as indigenous (primarily Wuitoto, Ticuna, and Murui). In contrast, only one donor from Leticia identified as indigenous, with most self-describing as *mestizos* (mixed-race). Most Km11 participants typically spend the day around their residence with high contact with livestock (82%), raising and sacrificing mostly poultry (chickens and ducks). Conversely, companion animal contact showed no significant difference between sites. Finally, over 90% of the donors stored water at home for domestic use.

**Table 1.**
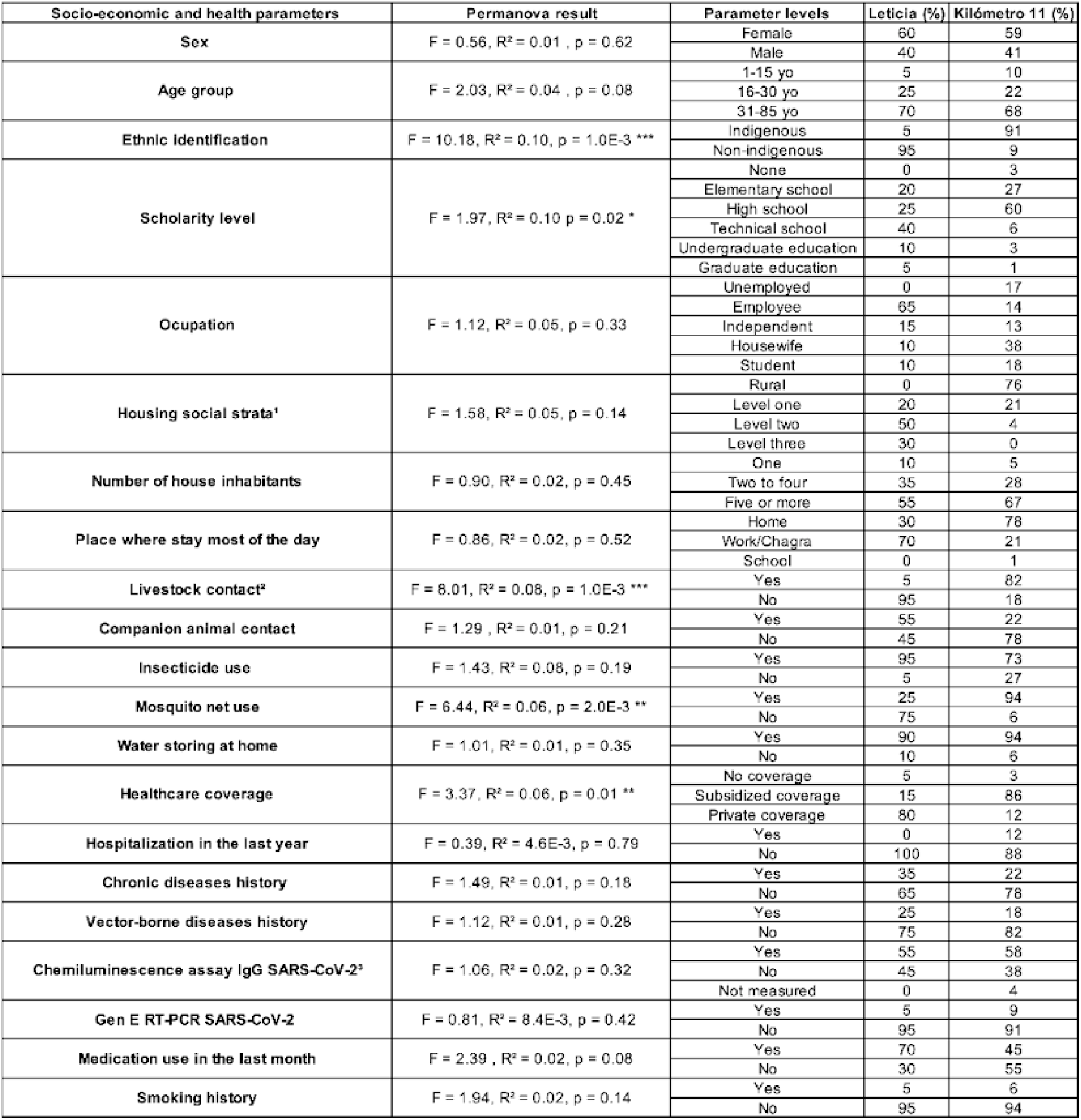
Socio-economic and health parameters for Leticia and Km11 groups. PERMANOVA results for each parameter with p-value indicating statically significant influence on microbiota structure. ¹Housing social strata based in the Colombian official strata divisions, rural and urban one to six levels. ²Livestock contact includes raising and/or sacrificing domestic animals. ³Three samples from Km11 were excluded because of the lack of biological material for testing.

Both groups reported low frequencies of historical diagnoses of vector-borne and chronic diseases (18-35%, with cardiovascular disease and hypertension being the most prevalent). Over 50% of donors at both locations had recent SARS-CoV-2 infection indicated by an IgG antibody assay, and less than 10% had an active positive infection based on RT-PCR (**Table 1**) although they were asymptomatic at the time of sampling. Medication use reported during the preceding month was high, particularly in Km11, especially for analgesics like paracetamol and ibuprofen.

To describe the bacterial microbiota compositions of the two groups, we sequenced the V4-V5 16S rRNA region. After preprocessing the reads, our final metabarcoding dataset comprised an average of 28,798 ± 15,564 sequences per sample, distributed across 466 OTUs. Most samples exhibited good taxonomic coverage, reaching the maximum number of OTUs with a subsampling of 10,000 sequences **(Fig. S1)**. Both richness (number of OTUs, p value = 0.0449) and diversity (Shannon index, p value = 0.001179) were statistically significantly reduced in the Leticia group **(Fig. 1A).** At the family level, *Prevotellaceae* was the most abundant group in the sampling, with >38% of the overall abundance for both locations **(Fig. 1B)**, followed by *Ruminococcaceae* (∼15%). We evaluated the influence of the surveyed variables on microbiota structure using a PERMANOVA analysis. The results revealed a statistically significant influence of location (F = 7.904, R² = 0.08; p = 0.002), with the Leticia samples clustering in the PCA plot (**Fig. 1C**). Other variables also differed between locations, such as ethnic identification, livestock contact, mosquito net use, healthcare coverage, and educational level, and these were significantly associated with the gut microbiota community structure (**Table 1**).

**Fig 1.**
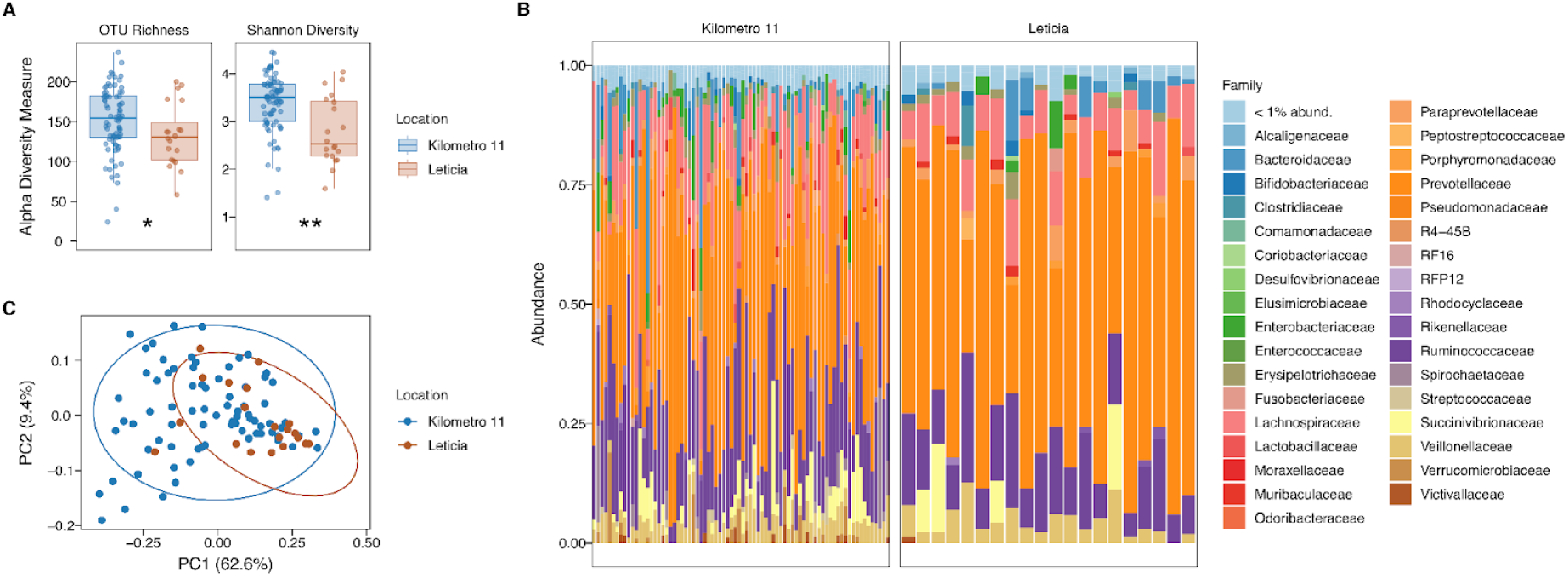
Leticia gut bacteria microbiota alpha and beta diversity analysis. **(A)** Alpha diversity estimators, (a) Number of OTUs (richness) and (b) Shannon Index (diversity). Difference between locations evaluated using Wilcoxon Paired-tests. *p = 0.05–0.005, **p = 0.0049–0.0005, ***p < 0.00049. **(B)** Relative bacterial family abundance for locations. Families with < 1% abundance were merged into one group. **(C)** PCA plot of bacterial community structure based on the weighted UniFrac distances.

Next, to identify the taxa driving differences between Leticia and Km11, we analyzed bacterial differential abundance using two approaches. For the Wald significance test, 135 out of 466 total OTUs showed a statistically significant log fold change (**Table S2**), with increased abundance in Km11 compared to Leticia (**Fig. 2A**). Most of these OTUs (55%) belong to the clostridial families *Ruminococcaceae* and *Lachnospiraceae*, and 22% were absent in Leticia samples (**Table S2**). The selected family abundance analysis (**Fig. 2B**) showed significant reductions of VANISH taxa in Leticia, with *Spirochaetaceae* locally extinct and only *Prevotellaceae* increased in the urban setting. No significant difference was observed for BloSSUM groups, being in low abundance for most donors.

**Fig 2.**
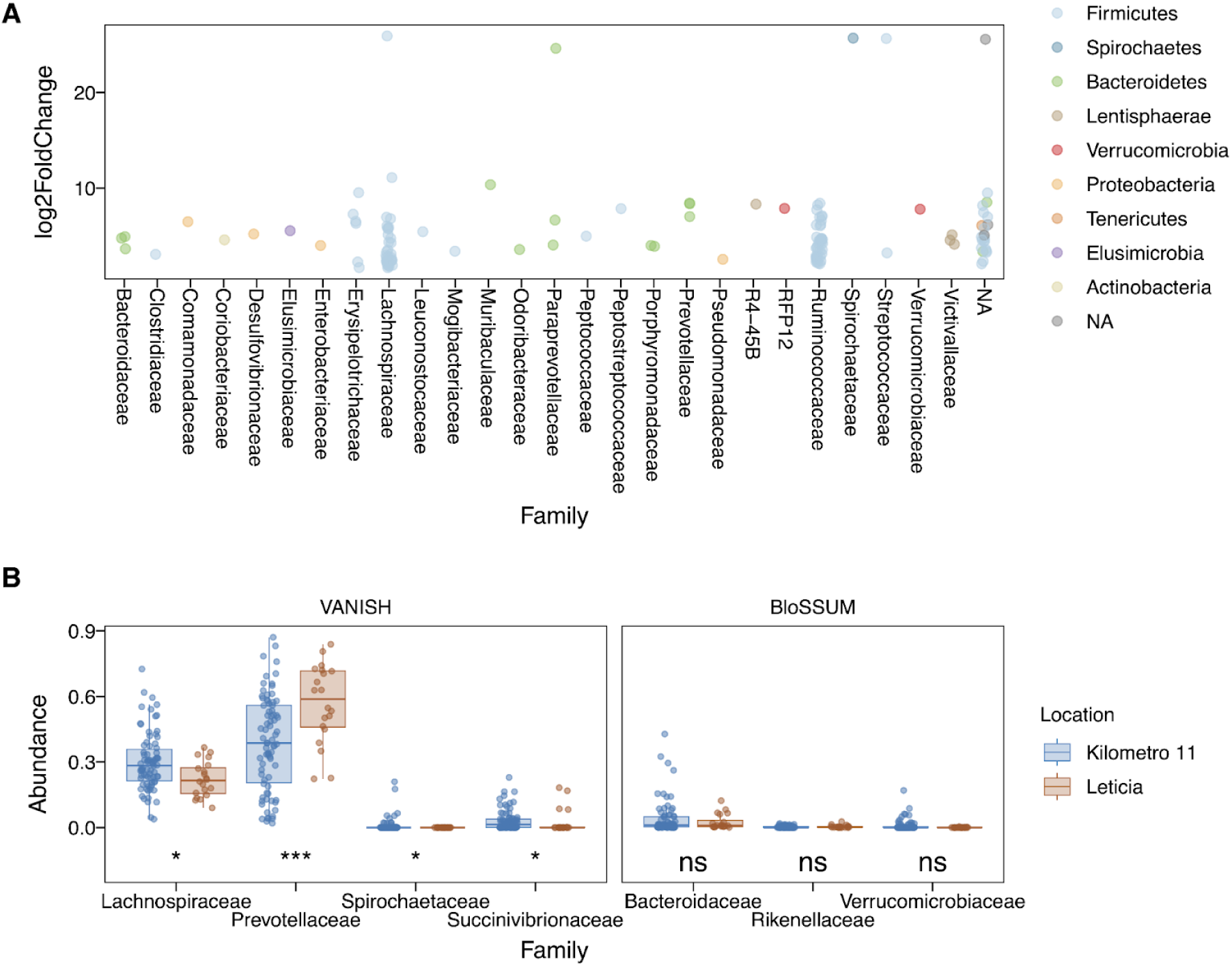
Differential abundance analysis between Leticia and Km11. **(A)** Significant OTUs according to the Wald significance test. Km11 as the treatment and Leticia as the control group, with an α = 0.01 threshold cutoff. **(B)** Differential abundance of VANISH and BloSSUM taxa. Difference between locations evaluated using Wilcoxon paired-tests. *p = 0.05–0.005, **p = 0.0049–0.0005, ***p < 0.00049.

In order to infer physiological implications of these bacterial microbial repertoires, we performed a predictive functional analysis finding a total of 331 predicted metabolic pathways. Using a Wald significance test, we identified 30 pathways with significant differences in relative abundance between the two locations (**Fig. 3** and **Table S3**). Compared to Leticia, 14 pathways were increased in Km11, with half of these belonging to fatty acid and lipid biosynthesis pathways. A total of 16 pathways were increased in Leticia, including pathways associated with aerobic respiration like cofactors biosynthesis pathways.

**Fig 3.**
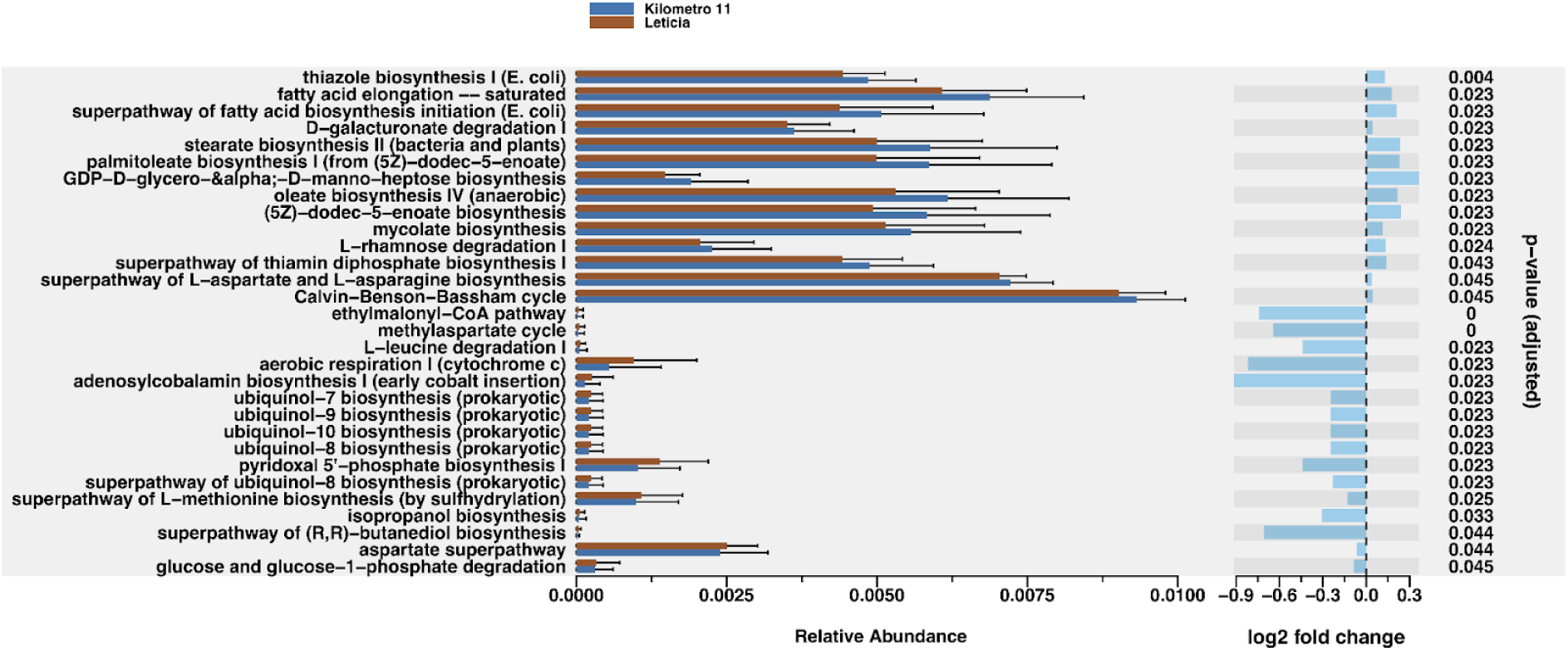
Predictive bacterial metabolic pathways analysis. **(Left)** Relative abundance of pathways with differential abundance between locations. **(Right)** Logarithmic fold change between pathways based on Wald Significance Test. Km11 as the treatment and Leticia as the control group, with an α = 0.01 threshold cutoff.

To evaluate the bacterial community compositions in a regional context, we performed a comparative gut bacteria microbiota analysis comprising nine datasets, including six Amazonian datasets (two urban groups including our sampling, and four rural groups) and three urban non-Amazonian datasets, for a total of 489 samples (**Table S1; Fig. S2**). For the alpha diversity (**Fig. 4A**), rural Amazonian groups showed higher richness than urban communities, where Leticia has similar values to the other Colombian urban microbiotas. We performed a PERMANOVA analysis to evaluate how much the sampled location can explain the microbiota structure (F = 37.94, R² = 38.09; p = 0.001). This was visualized in the PCA, where Ohio samples formed a separated cluster from three South American subgroups, Colombian urban Medellín-Bogotá, one transitional group for the Amazonian rural Tsimané, and the Leticia samples indistinct from Brazilian rural and urban populations (**Fig. 4B**). The clustering of Leticia and Brazilian samples is explained in the selected family analysis (**Fig. 4C**), where VANISH taxa *Prevotellaceae* and *Succinivibrionaceae* are increased compared to the other populations, while BloSSUM groups dominated in the urban populations (*Bacteroidaceae* and *Rikenellaceae* in Ohio, and *Verrucomicrobiacea* in Bogotá and Medellín) are reduced. Belém, the other Amazonian city evaluated, is distinguished from Leticia for the higher abundance of *Bacteroidaceae,* even with some samples clustering with the Ohio samples.

**Fig 4.**
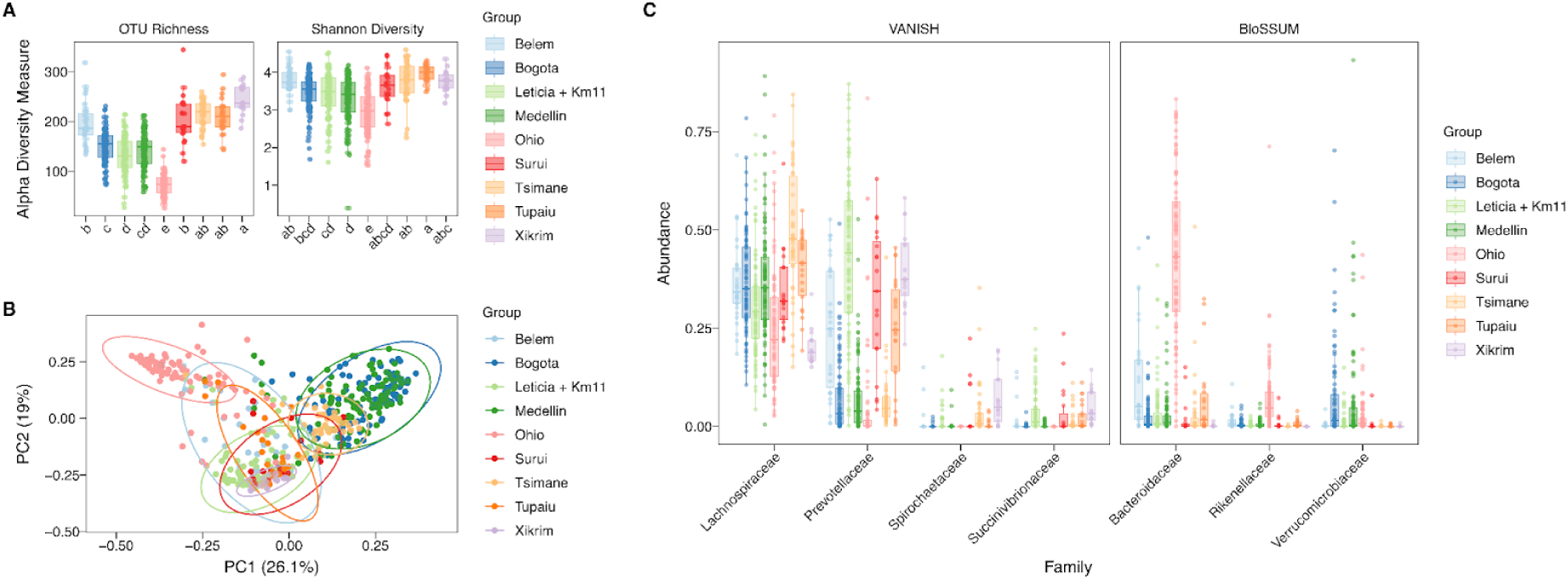
Amazonian and non-Amazonian gut bacteria microbiota datasets alpha and beta diversity comparative analysis. **(A)** Alpha diversity estimators of number of OTUs (richness) and Shannon Index (diversity). Letters reflect grouping and differences between datasets evaluated using Tukey HSD test. **(B)** PCA plot of bacterial community structure based on the weighted UniFrac distances. **(C)** Abundance of VANISH and BloSSUM taxa across bacteria microbiota datasets.

To describe the parasitic protists and nematodes associated with the Leticia gut microbiota, we performed an 18s rRNA analysis using the VESPA (Vertebrate Eukaryotic endoSymbiont and Parasite Analysis) protocol for eukaryotic endosymbiont metabarcoding [51]. Amplicons were generated targeting the 18S rRNA gene V4 region, sequenced, and recovered reads were classified into seven taxonomic categories using the PR2 reference sequence database. Parasitic protist sequences were the most common group, followed by human host. Parasitic nematodes were recovered in a low abundance (<1%) (**Fig. 5A**). Most individuals in both locations were infected with two or more parasitic protist taxa (**Fig. 5B**). Nematode infections were mostly detected in rural Km11 (**Fig. 5B**). Only two nematode taxa, *Enterobius* and *Necator*, were detected in urban Leticia, while seven genera were found in Km11 at rates ranging from 1 to 17% (**Fig. 5C**). Five protist taxa were detected in both locations, with *Blastocytis* being the most common, detected in >85% of samples from both locations (**Fig. 5C**).

**Fig 5.**
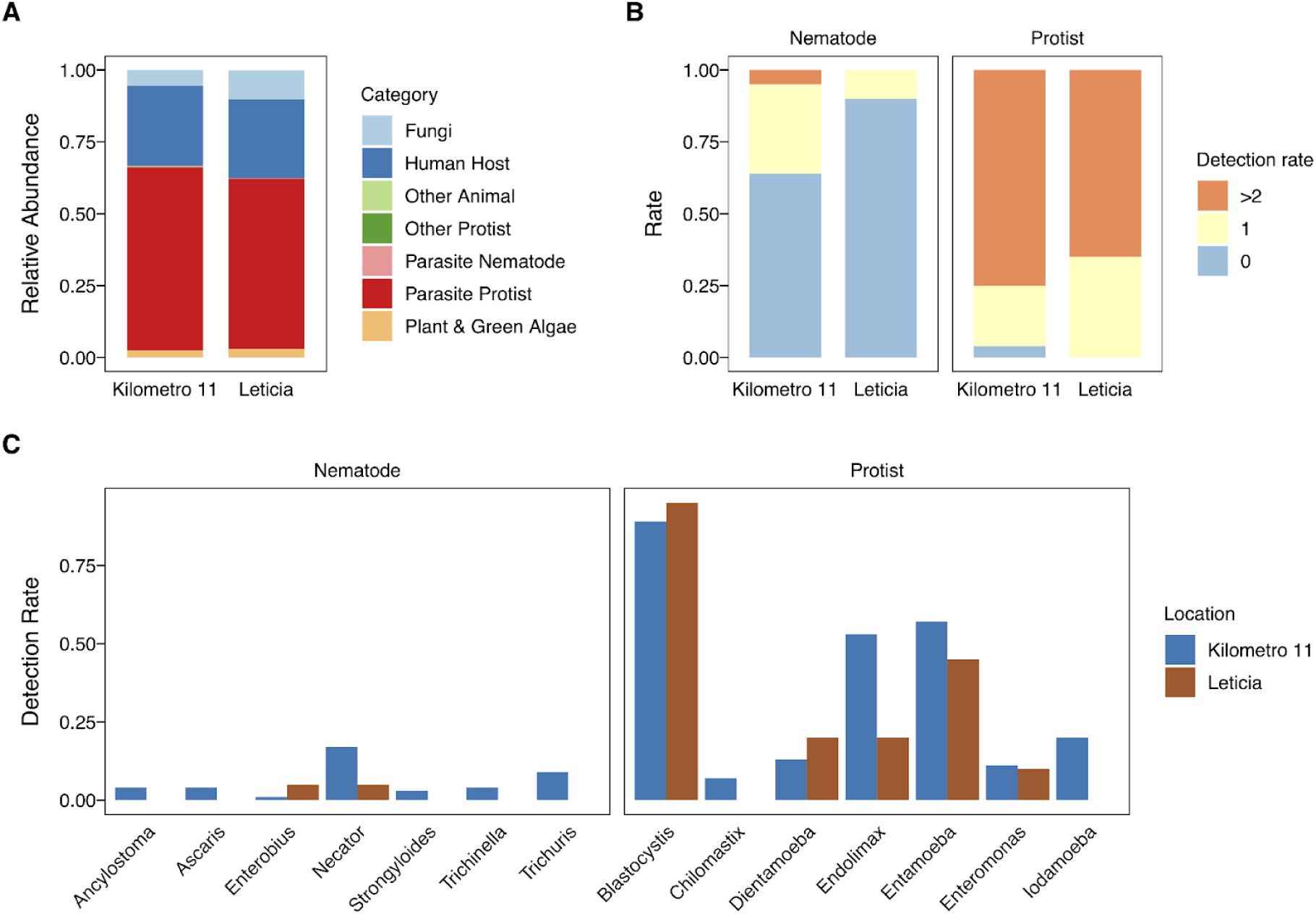
Leticia eukaryotic parasite metabarcoding description. **(A)** Relative sequence abundance from the 18s rRNA metabarcoding for each taxonomic category. **(B)** Overall detection rate of parasite nematodes and protists in Km11 and Leticia samples and **(C)** breakdown of taxa detected.

Given the extensive literature indicating that parasite infection influences the host microbial environment, we tested whether there was an association with protist or helminth infection and either bacterial diversity and abundance. PERMANOVA tests were used to evaluate alpha diversity metrics for the number of observed OTUs (richness), Shannon index (diversity), and abundance of *Prevotellaceae* and *Lachnospiraceae* family of bacteria. The Leticia and Km11 datasets were combined into one group, and comparisons were made between positive and negative infection status with any STH taxa or infection with any of the five most prevalent protists (*Blastocystis*, *Dientamoeba*, *Endolimax*, *Entamoeba*, and *Enteromonas*). Parasite infection was significantly associated with increased richness of bacterial tax for *Endolimax* (F = 12.137, R² = 0.115, p = 0.001) and *Entamoeba* (F = 19.834, R² = 0.176, p = 0.001). Infection with the following protist and nematode parasites were associated with increased diversity; *Endolimax* (F = 12.618, R² = 0.119, p = 0.001), *Entamoeba* (F = 12.625, R² = 0.119, p = 0.003), and STH infection (F = 6.919, R² = 0.069, p = 0.007). For the bacterial taxonomic abundance, STH infection influenced both *Prevotellaceae* (F = 6.722, R² = 0.068, p = 0.001) and *Lachnospiraceae* (F = 4.753, R² = 0.048, p = 0.018). All the results were confirmed with Wilcoxon paired tests comparing between positive and negative infection samples (**Fig. 6**).

**Fig 6.**
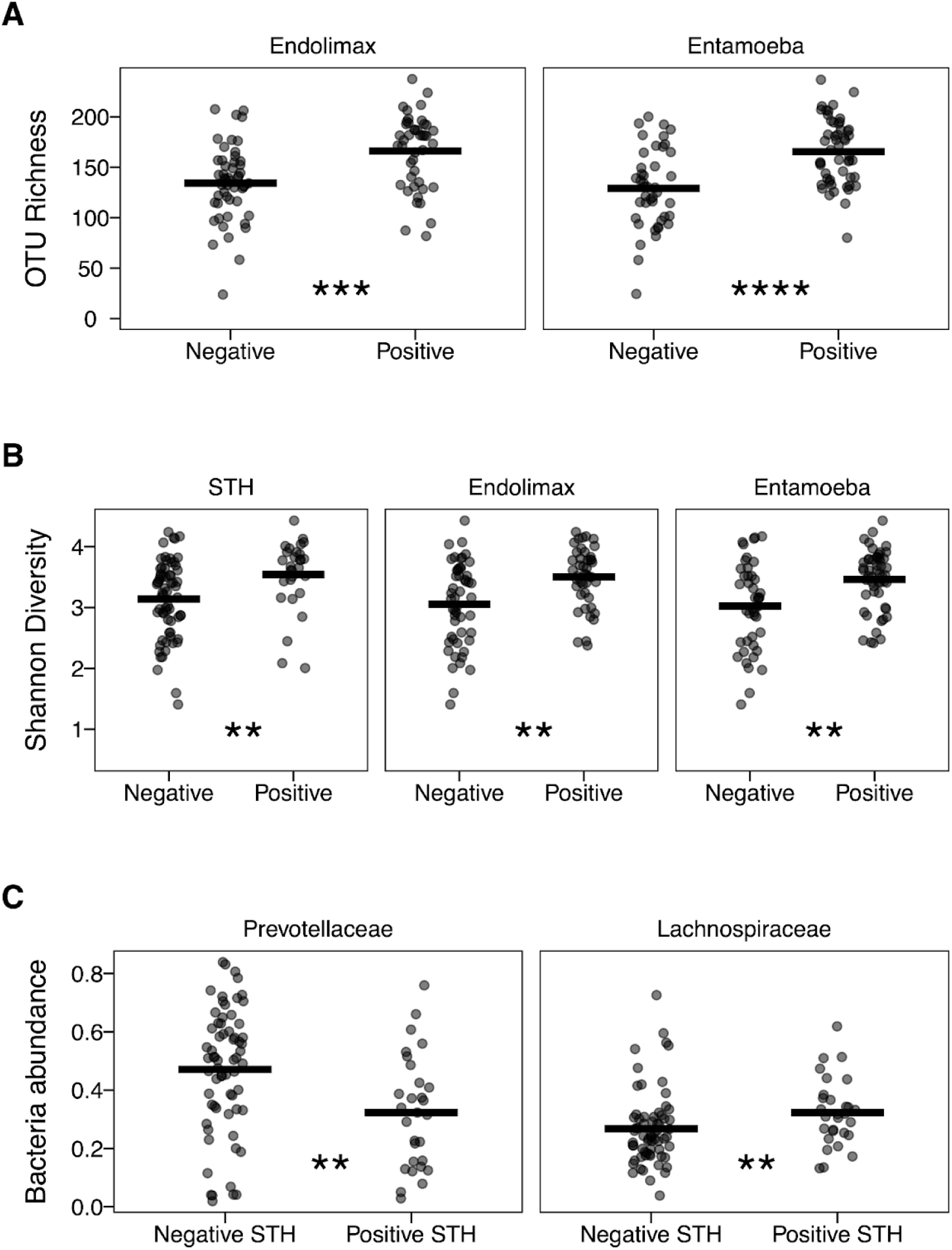
Differential analysis between parasite positive and negative Leticia (both urban and rural) samples. **(A)** Richness (observed number of OTUs) for *Endolimax* and *Entamoeba* infection. **(B)** Diversity (Shannon index) for STH, *Endolimax*, and *Entamoeba* infection. **(C)** *Prevotellaceae* and *Lachnospiraceae* abundance for STH infection. Difference between infection status evaluated using Wilcoxon paired-tests. *p = 0.05–0.005, **p = 0.0049–0.0005, ***p < 0.00049.

## Discussion

This study sheds new light on the gut bacterial microbiota of urban and rural populations in the Colombian Amazonian. In urban Leticia, we found lower bacterial diversity than in Km11 communities, reflecting reduced abundance of families related to non-processed foods (VANISH taxa), such as *Lachnospiraceae*, *Ruminococcaceae*, *Succinivibrio*, and *Spirochaetaceae*. The latter includes spirochetes of the genus *Treponema*, which are strongly associated with traditional rural populations of non-“Western” lifestyles [35]. Similar reductions in microbiota diversity have also been reported in other locations undergoing demographic and cultural transitions [53,54]. However, a noteworthy difference for Leticia is the increase of *Prevotella* instead of taxa associated with processed foods diets (BloSSUM taxa) in the urban setting.

Decreased microbial diversity in samples from the urban, mostly non-indigenous Leticia population compared to the rural, mostly indigenous Km11 population may be explained by lifestyle differences. Most of the Km11 donors practice activities such as small poultry farming for economic sustenance [55]. Outdoor activity [56] and livestock raising [57,58] have been shown to enrich microbiota diversity. Also, the diet in urban Leticia has changed significantly in recent decades compared to rural areas of the Amazonia, with reduced consumption of traditional non-industrialized foods like fish broth, wild animals, cassava-derived products like casabe and farinha, and fruit-based juices, and increased consumption of products such as packaged chicken, eggs, rice, canned foods, and powdered drink mixes of coffee, cocoa, or fruit [55].

Our comparative analyses show that the gut microbiota of people in and around Leticia is similar to that described within the Brazilian Amazonian urban and rural communities [4,6], with a high *Prevotellaceae* abundance and in general low presence of BloSSUM taxa. This pattern may be explained by similarities in the diet of these riverine communities along the Amazonian basin, which is still mostly composed of fish and polysaccharide-rich foods like cassava [4]. This differs from the lowland forest Tsimané community in Bolivia [34], the other Amazonian group evaluated, which has a diet rich in plant foraging and wild animals [59]. The taxonomic differences with Bogotá and Medellín, the other Colombian urban microbiotas, where BloSSUM taxa are more abundant, seem to reinforce the concept of a “tropical urban” category to describe microbiota of habitats of urban areas in tropical regions that are in different stages of microbiota “westernization” compared to non-tropical populations [4], although the different levels of urban transition, exemplified in the taxonomic differences between Leticia and Belém, indicate the necessity to further sample more Amazonian urban locations.

We also described the Leticia gut eukaryotic parasite community using a recently developed 18s rRNA metabarcoding protocol. We found a high abundance of intestinal parasites in both urban and rural samples, with *Endolimax* and *Entamoeba* presence having a positive effect on microbial richness and diversity. Infection with protists is already known to influence microbiota structure, being associated with higher microbial richness and abundance of VANISH taxa like *Prevotellaceae* and *Ruminococcaceae* [12,13]. We found that STH parasites are associated with higher bacterial microbial diversity and VANISH taxa abundance. A similar result, increased bacterial species diversity and *Prevotellaceae* abundance, was reported in a cohort of Colombian harboring mixed STH infections [60]. These results indicate potential parasite-microbiome interactions that could influence human health. This phenomenon has been extensively studied for both protists and helminths, which have been proposed to be beneficial by protecting against allergic and metabolic diseases [61–63].

Finally, predictive metabolic analysis indicates several significant results that may be relevant to health outcomes of the microbiota structure in the study area. Urban samples had relative depletion of fatty acid biosynthesis pathways. This finding may be explained by reduced abundance of *Lachnospiraceae*, which are key producers of butyrate and other short-chain fatty acids and decrease in diets high in saturated fatty acids [64]. Interestingly, depletion in short chain fatty acid producers was also observed in *Trichuris* infected individuals in a large study spanning Côte d’Ivoire, Laos, and Tanzania [65]. There was also an increase in the aerobic respiration pathways in urban samples, which may indicate increased saturated fatty acid consumption. These changes can increase the epithelial oxygenation in the colon, triggering a microbiome dysbiosis, characterized by an elevated abundance of facultatively aerobic bacteria compared to the healthy microbiota composition dominated by anaerobic bacteria [66]. Finally, although *Prevotellaceae* has been associated with a healthy microbiota, the high prevalence found in Leticia should be carefully interpreted, as this group has also been linked with inflammatory autoimmune diseases like rheumatoid arthritis [67].

Differences in the taxonomic composition and predicted physiology of the urban and rural Leticia microbiota documented in this study are likely associated with cultural and health transitions and intestinal protozoa and STH infections. These findings have relevance to public health, as such changes may underlie increases in chronic non-communicable diseases in the region, highlighting the need for further investigations into microbiota dynamics among urban and rural populations across the Colombian Amazon. This expanded analysis will be important for enhancing our understanding of the local health transitions and implementing proactive measures to improve public health outcomes and healthcare system preparedness.

## Data Availability

All relevant data are within the manuscript and its Supporting Information files. Sequencing information has been uploaded to NCBI SRA database (bioproject accession number PRJNA1246579), with the exception of reads mapping to the human genome. These were removed in the interests of protecting human subjects.

## Acknowledgments

This work was supported by funding from a UW-Madison Global Health Institute Pilot Grant (A.J.E.), NIH-NIAID funding R01AI151171 (M.Z.) and R21AI163592 (T.L.G.). S.D. was supported by a UW-Madison Global Health Institute Postdoctoral Fellowship and L.A.O. by the UW Madison Parasitology and Vector Biology Training Grant (T32AI007414).

---

**Table S1.** Sample list for Amazonian and non-Amazonian datasets used for the comparative gut bacterial microbiota analysis.

**Table S2.** Overall abundance and logarithmic fold change value for OTUs with a differential abundance between Leticia and Km11 based on the Wald Significance Test.

**Table S3**. Metacyc database classification of the metabolic pathways with a differential abundance between Leticia and Km11 based on the Wald Significance Test.

**Fig S1.**
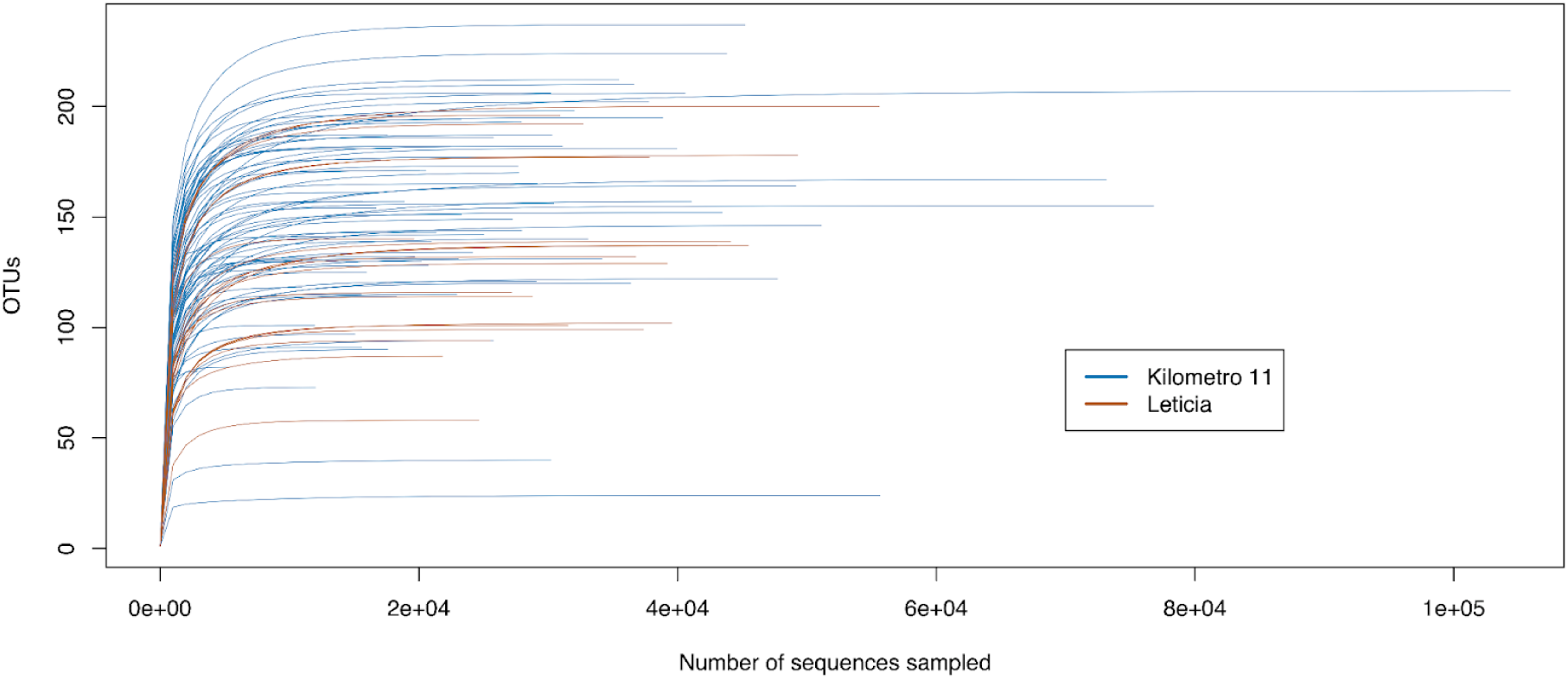
Rarefaction curve for Leticia and Km11 samples.

**Fig S2.**
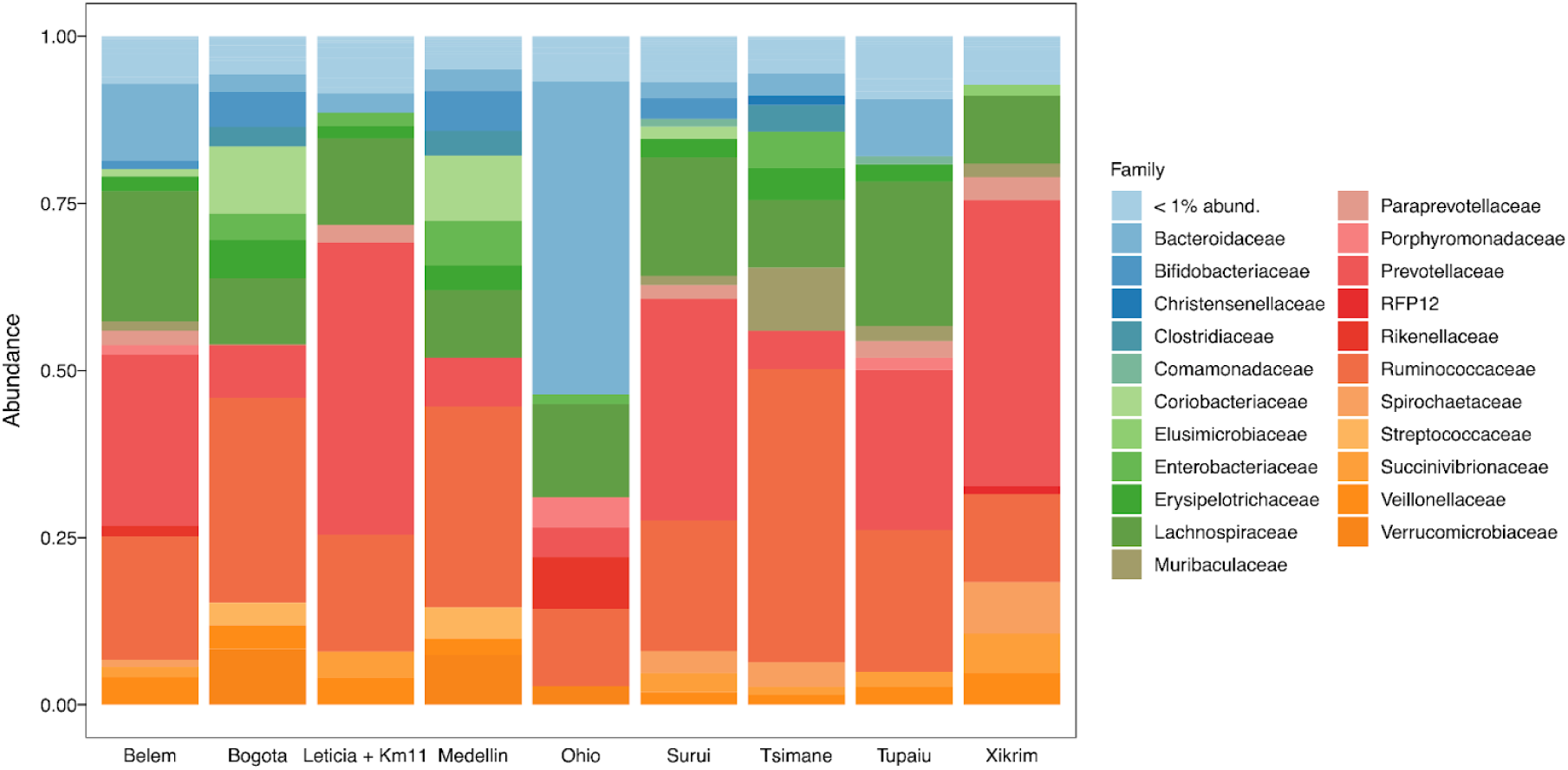
Relative bacterial family abundance for Amazonian and non-Amazonian datasets with each bar summarizing the total of samples per dataset. Families with < 1% abundance were merged into one group.

## References

1. Reynoso-García J, Miranda-Santiago AE, Meléndez-Vázquez NM, Acosta-Pagán K, Sánchez-Rosado M, Díaz-Rivera J, et al. A complete guide to human microbiomes: Body niches, transmission, development, dysbiosis, and restoration. Front Syst Biol. 2022;2. doi:10.3389/fsysb.2022.951403

2. Lokmer A, Aflalo S, Amougou N, Lafosse S, Froment A, Tabe FE, et al. Response of the human gut and saliva microbiome to urbanization in Cameroon. Sci Rep. 2020;10: 2856.

3. Soto-Girón MJ, Peña-Gonzalez A, Hatt JK, Montero L, Páez M, Ortega E, et al. Gut microbiome changes with acute diarrheal disease in urban versus rural settings in northern Ecuador. Am J Trop Med Hyg. 2021;104: 2275–2285.

4. Schaan AP, Sarquis D, Cavalcante GC, Magalhães L, Sacuena ERP, Costa J, et al. The structure of Brazilian Amazonian gut microbiomes in the process of urbanisation. NPJ Biofilms Microbiomes. 2021;7: 65.

5. Sun S, Wang H, Howard AG, Zhang J, Su C, Wang Z, et al. Loss of novel diversity in human gut Microbiota associated with ongoing urbanization in China. mSystems. 2022;7: e0020022.

6. Sonnenburg ED, Sonnenburg JL. The ancestral and industrialized gut microbiota and implications for human health. Nat Rev Microbiol. 2019;17: 383–390.

7. Sonnenburg JL, Sonnenburg ED. Vulnerability of the industrialized microbiota. Science. 2019;366: eaaw9255.

8. Urban population (% of total population). In: World Bank Open Data [Internet]. [cited 18 Mar 2025]. Available: https://data.worldbank.org/indicator/SP.URB.TOTL.IN.ZS

9. Burgess SL, Gilchrist CA, Lynn TC, Petri WA Jr. Parasitic protozoa and interactions with the host intestinal Microbiota. Infect Immun. 2017;85. doi:10.1128/IAI.00101-17

10. Llinás-Caballero K, Caraballo L. Helminths and bacterial Microbiota: The interactions of two of humans’ “old friends.” Int J Mol Sci. 2022;23: 13358.

11. Lee SC, Tang MS, Lim YAL, Choy SH, Kurtz ZD, Cox LM, et al. Helminth colonization is associated with increased diversity of the gut microbiota. PLoS Negl Trop Dis. 2014;8: e2880.

12. Alzate JF, Toro-Londoño M, Cabarcas F, Garcia-Montoya G, Galvan-Diaz A. Contrasting microbiota profiles observed in children carrying either Blastocystis spp. or the commensal amoebas Entamoeba coli or Endolimax nana. Sci Rep. 2020;10: 15354.

13. Even G, Lokmer A, Rodrigues J, Audebert C, Viscogliosi E, Ségurel L, et al. Changes in the Human Gut Microbiota Associated With Colonization by Blastocystis sp. and Entamoeba spp. in Non-Industrialized Populations. Front Cell Infect Microbiol. 2021;11: 533528.

14. Rosa BA, Supali T, Gankpala L, Djuardi Y, Sartono E, Zhou Y, et al. Differential human gut microbiome assemblages during soil-transmitted helminth infections in Indonesia and Liberia. Microbiome. 2018;6: 33.

15. Kupritz J, Angelova A, Nutman TB, Gazzinelli-Guimaraes PH. Helminth-induced human gastrointestinal dysbiosis: A systematic review and meta-analysis reveals insights into altered taxon diversity and microbial gradient collapse. MBio. 2021;12: e0289021.

16. Tee MZ, Er YX, Easton AV, Yap NJ, Lee IL, Devlin J, et al. Gut microbiome of helminth-infected indigenous Malaysians is context dependent. Microbiome. 2022;10: 214.

17. Stensvold CR, van der Giezen M. Associations between gut Microbiota and common luminal intestinal parasites. Trends Parasitol. 2018;34: 369–377.

18. Morton ER, Lynch J, Froment A, Lafosse S, Heyer E, Przeworski M, et al. Variation in rural African gut Microbiota is strongly correlated with colonization by Entamoeba and subsistence. PLoS Genet. 2015;11: e1005658.

19. Carey MA, Medlock GL, Alam M, Kabir M, Uddin MJ, Nayak U, et al. Megasphaera in the stool Microbiota is negatively associated with diarrheal cryptosporidiosis. Clin Infect Dis. 2021;73: e1242–e1251.

20. Brosschot TP, Reynolds LA. The impact of a helminth-modified microbiome on host immunity. Mucosal Immunol. 2018;11: 1039–1046.

21. Gause WC, Maizels RM. Macrobiota - helminths as active participants and partners of the microbiota in host intestinal homeostasis. Curr Opin Microbiol. 2016;32: 14–18.

22. McFarlane AJ, McSorley HJ, Davidson DJ, Fitch PM, Errington C, Mackenzie KJ, et al. Enteric helminth-induced type I interferon signaling protects against pulmonary virus infection through interaction with the microbiota. J Allergy Clin Immunol. 2017;140: 1068–1078.e6.

23. Ramanan D, Bowcutt R, Lee SC, Tang MS, Kurtz ZD, Ding Y, et al. Helminth infection promotes colonization resistance via type 2 immunity. Science. 2016;352: 608–612.

24. Easton AV, Quiñones M, Vujkovic-Cvijin I, Oliveira RG, Kepha S, Odiere MR, et al. The impact of anthelmintic treatment on human gut Microbiota based on cross-sectional and pre- and postdeworming comparisons in western Kenya. MBio. 2019;10. doi:10.1128/mBio.00519-19

25. Gerbe F, Sidot E, Smyth DJ, Ohmoto M, Matsumoto I, Dardalhon V, et al. Intestinal epithelial tuft cells initiate type 2 mucosal immunity to helminth parasites. Nature. 2016;529: 226–230.

26. Howitt MR, Lavoie S, Michaud M, Blum AM, Tran SV, Weinstock JV, et al. Tuft cells, taste-chemosensory cells, orchestrate parasite type 2 immunity in the gut. Science. 2016;351: 1329–1333.

27. Schneider C, O’Leary CE, von Moltke J, Liang H-E, Ang QY, Turnbaugh PJ, et al. A metabolite-triggered tuft cell-ILC2 circuit drives small intestinal remodeling. Cell. 2018;174: 271–284.e14.

28. Fung C, Fraser LM, Barrón GM, Gologorsky MB, Atkinson SN, Gerrick ER, et al. Tuft cells mediate commensal remodeling of the small intestinal antimicrobial landscape. Proc Natl Acad Sci U S A. 2023;120: e2216908120.

29. Ndjim M, Gasmi I, Herbert F, Joséphine C, Bas J, Lamrani A, et al. Tuft cell acetylcholine is released into the gut lumen to promote anti-helminth immunity. Immunity. 2024;57: 1260–1273.e7.

30. Miguel Alexiades DP. La Urbanización Indígena En La, Amazonia; Un nuevo contexto de articulación social y territorial. Gazeta de Antropologia. 2016;32.

31. Obregon-Tito AJ, Tito RY, Metcalf J, Sankaranarayanan K, Clemente JC, Ursell LK, et al. Subsistence strategies in traditional societies distinguish gut microbiomes. Nat Commun. 2015;6: 6505.

32. Clemente JC, Pehrsson EC, Blaser MJ, Sandhu K, Gao Z, Wang B, et al. The microbiome of uncontacted Amerindians. Sci Adv. 2015;1. doi:10.1126/sciadv.1500183

33. Yatsunenko T, Rey FE, Manary MJ, Trehan I, Dominguez-Bello MG, Contreras M, et al. Human gut microbiome viewed across age and geography. Nature. 2012;486: 222–227.

34. Sprockett DD, Martin M, Costello EK, Burns AR, Holmes SP, Gurven MD, et al. Microbiota assembly, structure, and dynamics among Tsimane horticulturalists of the Bolivian Amazon. Nat Commun. 2020;11: 3772.

35. Angelakis E, Bachar D, Yasir M, Musso D, Djossou F, Gaborit B, et al. Treponema species enrich the gut microbiota of traditional rural populations but are absent from urban individuals. New Microbes New Infect. 2019;27: 14–21.

36. Alencar RM, Martínez JG, Machado VN, Alzate JF, Ortiz-Ojeda CP, Matias RR, et al. Preliminary profile of the gut microbiota from amerindians in the Brazilian amazon experiencing a process of transition to urbanization. Braz J Microbiol. 2024;55: 2345–2354.

37. Aponte Motta J, Rojas Correal A, de Carvalho Coutinho T. Alimentando la ciudad y resistiéndola: los pueblos indígenas en el complejo urbano transfronterizo entre Brasil, Colombia y Perú en la Amazonia. Lat Am Caribb Ethn Stud. 2023; 1–24.

38. National Administrative Department of Statistics (DANE). PROYECCIONES DE POBLACIÓN. [cited Mar 2025]. Available: https://www.dane.gov.co/index.php/estadisticas-por-tema/demografia-y-poblacion/proyecciones-de-poblacion

39. Tobón MA. La fórmula biodiversidad - cultura y el poder político en el extremo sur del Trapecio Amazónico colombiano. Univ Humaníst. July/Dec 2006.

40. Cardona A, Villegas S, Lopez MS, Maya MA, Ortiz-Restrepo C, Hernandez-Ortiz O, et al. SARS-CoV-2 vaccinated breakthrough infections with fatal and critical outcomes in the department of Antioquia, Colombia. Research Square. 2022. doi:10.21203/rs.3.rs-963938/v1

41. Bolyen E, Rideout JR, Dillon MR, Bokulich NA, Abnet CC, Al-Ghalith GA, et al. Reproducible, interactive, scalable and extensible microbiome data science using QIIME 2. Nat Biotechnol. 2019;37: 852–857.

42. Callahan BJ, McMurdie PJ, Rosen MJ, Han AW, Johnson AJA, Holmes SP. DADA2: High-resolution sample inference from Illumina amplicon data. Nat Methods. 2016;13: 581–583.

43. Bokulich NA, Kaehler BD, Rideout JR, Dillon M, Bolyen E, Knight R, et al. Optimizing taxonomic classification of marker-gene amplicon sequences with QIIME 2’s q2-feature-classifier plugin. Microbiome. 2018;6: 90.

44. DeSantis TZ, Hugenholtz P, Larsen N, Rojas M, Brodie EL, Keller K, et al. Greengenes, a chimera-checked 16S rRNA gene database and workbench compatible with ARB. Appl Environ Microbiol. 2006;72: 5069–5072.

45. McMurdie PJ, Holmes S. phyloseq: an R package for reproducible interactive analysis and graphics of microbiome census data. PLoS One. 2013;8: e61217.

46. Dixon P. VEGAN, a package of R functions for community ecology. J Veg Sci. 2003;14: 927–930.

47. Love MI, Huber W, Anders S. Moderated estimation of fold change and dispersion for RNA-seq data with DESeq2. Genome Biol. 2014;15: 550.

48. Douglas GM, Maffei VJ, Zaneveld JR, Yurgel SN, Brown JR, Taylor CM, et al. PICRUSt2 for prediction of metagenome functions. Nat Biotechnol. 2020;38: 685–688.

49. Caspi R, Billington R, Keseler IM, Kothari A, Krummenacker M, Midford PE, et al. The MetaCyc database of metabolic pathways and enzymes - a 2019 update. Nucleic Acids Res. 2020;48: D445–D453.

50. Yang C, Mai J, Cao X, Burberry A, Cominelli F, Zhang L. ggpicrust2: an R package for PICRUSt2 predicted functional profile analysis and visualization. Bioinformatics. 2023;39. doi:10.1093/bioinformatics/btad470

51. Owens LA, Friant S, Martorelli Di Genova B, Knoll LJ, Contreras M, Noya-Alarcon O, et al. VESPA: an optimized protocol for accurate metabarcoding-based characterization of vertebrate eukaryotic endosymbiont and parasite assemblages. Nat Commun. 2024;15: 402.

52. Guillou L, Bachar D, Audic S, Bass D, Berney C, Bittner L, et al. The Protist Ribosomal Reference database (PR2): a catalog of unicellular eukaryote small sub-unit rRNA sequences with curated taxonomy. Nucleic Acids Res. 2013;41: D597–604.

53. Tamburini FB, Maghini D, Oduaran OH, Brewster R, Hulley MR, Sahibdeen V, et al. Short- and long-read metagenomics of urban and rural South African gut microbiomes reveal a transitional composition and undescribed taxa. Nat Commun. 2022;13: 926.

54. van der Vossen EWJ, Davids M, Bresser LRF, Galenkamp H, van den Born B-JH, Zwinderman AH, et al. Gut microbiome transitions across generations in different ethnicities in an urban setting-the HELIUS study. Microbiome. 2023;11: 99.

55. Peña-Venegas CP, Valderrama AM, Muñoz LEA, Rúa MNP. Seguridad alimentaria en comunidades indígenas del Amazonas: ayer y hoy. Instituto Amazónico de Investigaciones Científicas “SINCHI”; 2009.

56. Brame JE, Liddicoat C, Abbott CA, Breed MF. The potential of outdoor environments to supply beneficial butyrate-producing bacteria to humans. Sci Total Environ. 2021;777: 146063.

57. Moor J, Wüthrich T, Aebi S, Mostacci N, Overesch G, Oppliger A, et al. Influence of pig farming on human Gut Microbiota: role of airborne microbial communities. Gut Microbes. 2021;13: 1–13.

58. Sudatip D, Mostacci N, Thamlikitkul V, Oppliger A, Hilty M. Influence of occupational exposure to pigs or chickens on human gut microbiota composition in Thailand. One Health. 2022;15: 100463.

59. Kaplan H, Thompson RC, Trumble BC, Wann LS, Allam AH, Beheim B, et al. Coronary atherosclerosis in indigenous South American Tsimane: a cross-sectional cohort study. Lancet. 2017;389: 1730–1739.

60. Toro-Londono MA, Bedoya-Urrego K, Garcia-Montoya GM, Galvan-Diaz AL, Alzate JF. Intestinal parasitic infection alters bacterial gut microbiota in children. PeerJ. 2019;7: e6200.

61. Erb KJ. Can helminths or helminth-derived products be used in humans to prevent or treat allergic diseases? Trends Immunol. 2009;30: 75–82.

62. Chabé M, Lokmer A, Ségurel L. Gut protozoa: Friends or foes of the human gut Microbiota? Trends Parasitol. 2017;33: 925–934.

63. Piperni E, Nguyen LH, Manghi P, Kim H, Pasolli E, Andreu-Sánchez S, et al. Intestinal Blastocystis is linked to healthier diets and more favorable cardiometabolic outcomes in 56,989 individuals from 32 countries. Cell. 2024;187: 4554–4570.e18.

64. Telle-Hansen VH, Gaundal L, Bastani N, Rud I, Byfuglien MG, Gjøvaag T, et al. Replacing saturated fatty acids with polyunsaturated fatty acids increases the abundance of Lachnospiraceae and is associated with reduced total cholesterol levels-a randomized controlled trial in healthy individuals. Lipids Health Dis. 2022;21: 92.

65. Schneeberger PHH, Dommann J, Rahman N, Hürlimann E, Sayasone S, Ali S, et al. Profound taxonomic and functional gut microbiota alterations associated with trichuriasis: cross-country and country-specific patterns. bioRxiv. 2025. doi:10.1101/2025.03.21.641387

66. Lee J-Y, Tsolis RM, Bäumler AJ. The microbiome and gut homeostasis. Science. 2022;377: eabp9960.

67. Tett A, Pasolli E, Masetti G, Ercolini D, Segata N. Prevotella diversity, niches and interactions with the human host. Nat Rev Microbiol. 2021;19: 585–599.

